# “Backed into a Corner”: Lived experiences of receiving and providing involuntary psychiatric treatment under British Columbia’s Mental Health Act

**DOI:** 10.1101/2025.08.01.25332770

**Authors:** Mary Elizabeth Snow, Amy Salmon, Jenyo Banjo, Marina Morrow, Colleen Varcoe

## Abstract

British Columbia’s Mental Health Act permits the involuntarily detention and treatment of individuals who meet specific criteria. Over the past 15 years, British Columbia has seen an increasing trend in the number of involuntary psychiatric admissions. This qualitative study explores the lived experiences of people receiving and providing involuntary psychiatric treatment within two health organizations in British Columbia, Canada. Five focus groups were conducted with 23 individuals who had previously received involuntary psychiatric treatment at a facility operated by one of the two health organizations. All sessions were facilitated by individuals with lived experience of involuntary psychiatric treatment. Additionally, semi-structured interviews were conducted with 11 clinical staff and 10 non-clinical support personnel involved in delivering involuntary psychiatric treatment.

Data were analyzed using a thematic approach. Seven key themes were generated, including the limited availability of voluntary care options, the compounding role of social determinants of health in mental health crises, the lack of conclusive evidence supporting involuntary psychiatric treatment, the negative impacts on both patients and providers, and the critical role of peer support. These findings underscore the need for systemic reform to reduce reliance on coercive practices and to expand access to voluntary, community-based mental health supports that address underlying social and structural factors contributing to mental health crises.

## Introduction

British Columbia’s Mental Health (MH) Act allows a person to be detained and treated without consent, including situations in which a person cannot articulate consent and in situations where a person explicitly refuses treatment — even when they are capable of making their own decisions (1). A person can be involuntarily admitted under the authority of the MH Act to a designated facility if they meet the following criteria, known as the “involuntary admission criteria”; the person:

1. “is suffering from a mental disorder that seriously impairs the person’s ability to react appropriately to his or her environment or to associate with others”
2. “requires treatment in or through a designated facility”,
3. “requires care, supervision and control in or through a designated facility to prevent the person’s or patient’s substantial mental or physical deterioration or for the protection of the person or patient or the protection of others”, and
4. “cannot suitably be admitted as a voluntary patient.”(2)

Unlike most other jurisdictions in Canada, the BC MH Act operates on a deemed consent model which indicates that once a person is detained involuntarily, they are deemed to have consented to any psychiatric treatment that follows (3). The language of “in or through a designated facility” enables psychiatric treatment to be administered without a patient’s or substitute decision-maker’s consent either at a designated hospital on an in-patient basis (where the patient is detained), or in a community setting, for example, when treatment is delivered by Assertive Community Treatment teams (4,5). Those treated involuntarily in community settings under the MH Act are referred to as being on “extended leave” from a designated facility to which they can be forcibly recalled and detained, if they do not comply with the leave conditions.

The MH Act stipulates that involuntary admission status is to be used until the individual “improves to the point that they can continue as voluntary patients or resume their lives in the community.” (2). To prevent unnecessary and indefinite detention, patients are entitled to periodic review of their detention by an independent panel (1). However, unlike many other jurisdictions, British Columbia places the burden of requesting a review on patients who may not be fully aware of this right, or be comfortable and well equipped to make such requests (6). Furthermore, once detained for treatment under the MH Act, there are no legal or formal procedural mechanisms available to patients to ensure oversight or review of the safety or efficacy of the psychiatric treatment administered to them, nor is there a legal mechanism for detainees to challenge psychiatric treatment administered to them against their will (7). The only option available to patients who are concerned with their psychiatric treatment is to request a second medical opinion on the treatment’s appropriateness. There is no time limit for the opinion to be completed and the challenged psychiatric treatment may continue while the opinion is arranged (6). Moreover, the original physician is not required to change treatment based on the second opinion.

As has been noted by BC’s Office of the Ombudsperson, the power to limit one’s liberty, bodily autonomy, privacy, and other human rights are extraordinary state powers that must be exercised rarely and with exceptional caution (8). The province of BC currently has the highest rate of involuntary hospitalization for mental health and substance use issues in Canada (9) The rates of involuntary psychiatric admission and treatment have doubled in the last 15 years with over 20,000 people detained annually under the MH Act in BC between 2016 an 2021 (10). During the same period, BC saw an eight-fold increase in the number of people on extended leave i.e., receiving mandated treatment in the community (6), while a study drawing on two comprehensive systematic reviews concluded that there is no evidence of patient benefit from such practices, casting doubt over both the usefulness and ethics of these orders (11). When population growth is considered, the use of voluntary psychiatric admissions has significantly decreased on a per capita basis (8).

Some observers have noted a correlation between increases in involuntary detainment of individuals in psychiatric crisis and limited or non-existent access to voluntary mental health treatment options, including upstream services that may prevent psychiatric crises (12–15). Substance use disorders represented the third largest diagnosis type for people detained and involuntarily treated under the MH Act (14). This occurs despite the fact that decades of research have failed to provide conclusive and consistent evidence supporting the effectiveness of involuntary treatments for substance use issues (16,17). Negative outcomes have been observed pertaining to involuntary treatment of substance use matters, including increased risk of post-release mortality (18), overdose (19), interruptions to or limited access to opioid agonist treatment (20), negative effects on therapeutic relationships, exposure to further coercive measures (including restraints), as well as disproportionate harms to specific populations (21,22). Recent studies from BC have found that people who use drugs do not endorse the use of involuntary treatment, pointing instead to significant changes required to address shortcomings of the voluntary care system, including needs for expanded access to existing services and a wider range of harm reduction, early intervention, and treatment options (23,24).

A 2022 review by the BC Office of the Ombudsperson found that none of the health authorities in the province were fully compliant with the required documentation practices for involuntary psychiatric admissions under the MH Act (10). To document medico-legal compliance, the MH Act mandates specific forms to be completed by health professionals at prescribed times. BC Office of the Ombudsperson noted:

> For patients and their families, the lack of adequate documentation naturally raises questions about the reasons for detention. For example, could less restrictive alternatives have been used? Ultimately, the lack of documentation raises questions about whether individuals at their most vulnerable have been detained lawfully and fairly. As a result, public confidence in the system at large is jeopardized. (8)

In response, two BC health organizations undertook system-wide efforts to ensure documentation practices conform to the legal requirements for the involuntary admissions process under the MH Act, as well as efforts to improve care providers’ and patients’ understanding of the rights of people hospitalized involuntarily under the Act. At the same time, leaders and advocates within and outside these organizations recognized that demonstrating legal administrative compliance with the MH Act alone was not sufficient to understand whether or not both organizations were delivering equity-oriented care informed by intersectional analysis of health disparities. We understand equity-oriented care as services that are experienced by recipients as safe, destigmatizing, trauma– and violence-informed, culturally-safe, harm reduction-oriented, anti-racist, and gender-affirming (25,26).

Between 2021-2023, we undertook a participatory qualitative study to better understand the lived experiences of receiving and providing involuntary treatment in these organizations’ hospitals. Our aims were: (1) to document and examine, from an equity-oriented perspective, the experiences of receiving or providing involuntary psychiatric admission and treatment under the BC MH Act and (2) identify opportunities to improve care delivery during periods of psychiatric crises that align with principles of equity-oriented care.

## Methods

All aspects of the study were guided by an Advisory Committee comprised of people with lived or living experience (PWLLE) of receiving involuntary psychiatric treatment, clinicians who have provided involuntary psychiatric treatment, staff in community agencies supporting those receiving involuntary psychiatric treatment, clinicians and leaders working within the health organizations’ mental health and substance use services unit, and researchers with expertise in equity-oriented mental health policy and practice. This study was approved by the UBC– Providence Health Care Research Institute Research Ethics Board (REB #H22-02037).

Qualitative data were collected via interviews and focus groups with people who have (1) received treatment under the MH Act at the two health organizations’ facilities, (2) provided treatment under the MH Act, or (3) provided legal or peer support to those who have received such treatment. Methods such as in-depth interviews and focus groups allow for rich, detailed assessment of people’s everyday experiences, providing context that is essential for crafting strategies that truly meet the needs of these populations (27,28). Qualitative methods were prioritized for the ability to identify, from the perspective of individuals, how lived experiences are intertwined with systemic issues that contribute to health disparities, such as implicit bias in care delivery, cultural barriers to accessing services, and other factors that influence how different individuals and population groups interact with the healthcare system (29–32).

Furthermore, qualitative research fosters a deeper understanding of cultural contexts, which is essential when working with a diverse range of populations. It allows researchers to examine how factors such as personal or cultural beliefs, health-related practices, and individual and collective values influence health behaviours and perceptions of care (33–35). These insights are critical for developing interventions and policies that address the root causes of health inequities (36,37). This is particularly important for policymaking which aims to ensure that healthcare services are anti-racist, culturally safe, destigmatizing, trauma– and violence-informed, and responsive to the needs of all communities (26,38–41).

## Data Collection

### Focus groups

Advisory Committee members with lived experience of involuntary psychiatric admission and treatment strongly recommended that qualitative data collection with people who have received such treatment would best be conducted in a focus group setting. Participants were recruited between February 1 and April 30, 2023 through social media and community agencies that work with PWLLE of involuntary psychiatric admission and treatment. Interested individuals were asked to contact a member of the research team for eligibility screening. Eligible participants had to be 19 years of age or older and have a history of involuntary psychiatric admission and treatment at any of the facilities operated by the two health organizations within the past five years.

To accommodate participants’ preferences, two focus groups were held in person and three were conducted virtually. Because these organizations operate healthcare facilities in urban and rural settings, one of the virtual focus groups was specifically for participants from rural communities. Attendance at the focus groups ranged from 2 to 5 participants and a total of 23 people participated. All participants provided informed written consent to participate in the study; however, only 6 participants consented to provide information about their gender or age. Of these 6 participants who provided demographic information, 4 (66.7%) identified as female. The mean age was 49.5 years (SD = 15.6; range: 25-63 years). One participant (16.7%) self-identified as Indigenous.

Focus groups were designed to minimize re-traumatization, prioritize participant agency, and acknowledge structural violence embedded in mental health systems. Thus, in-person focus groups were held in community locations that were not healthcare facilities. Private space outside the focus group room was available to allow in-person participants to step away from the group for some time if needed. Breakout rooms were made available during virtual focus groups for the same purpose. In both the in-person and virtual breakout rooms, a research team member was available to provide emotional and social support if requested. All focus groups were facilitated by one or two PWLLE with support from two research team members with qualitative research experience. Focus groups lasted 1.5 to 2 hours each, were audio recorded and transcribed verbatim.

### Interviews

Advisory Committee members from health professions recommended that semi-structured interviews with those who provide treatment or support to individuals who have received admission and treatment under the MH Act be conducted individually. Participants were recruited between February 1 and April 30, 2023 through purposive sampling of individuals who provide services to people admitted for involuntary psychiatric treatment under the MH Act. Participants were strategically selected based on their professional roles to ensure diverse representation. Interview participants provided informed written consent.

Two researchers with qualitative interview experience (AS and MES) conducted interviews with 21 individuals. Table 1 shows participants’ roles. Interviews lasted approximately 1 hour each, were conducted virtually, audio recorded, and transcribed verbatim.

**Table 1.** Categories of participants who provide clinical care and non-clinical support.

| Category | Role | Number of participants |
| --- | --- | --- |
| Clinician | Nurse | 3 |
|  | Physician | 5 |
|  | Social Worker | 3 |
| Service Provider<sup>a</sup> | Lawyer/Legal Advocate | 4 |
|  | Peer Support Worker | 4 |
|  | Program Leader/Manager | 2 |

*Note.* <sup>a</sup> We use the term “service provider” to refer to professionals providing non-clinical support for individuals admitted and treated involuntarily under the BC MH Act.

### Data Analysis

Data were analyzed using a thematic analytic approach using NVivo (version 14) to organize the data and track the analysis. Analysis of qualitative data from individual interviews and focus groups followed the steps articulated by Braun and Clarke (42,43) for conducting reflexive thematic analysis:

1. Familiarization. Multiple research team members, including those who conducted interviews and those who had not participated in the data collection process, thoroughly read and re-read the data, and then began identifying initial concepts or patterns generated in the narrative accounts.
2. Coding. Two members systematically and independently reviewed transcripts and reflexive notes generated in the familiarization state to generate codes for segments relevant to the study aims. They held multiple iterative meetings to discuss coding discrepancies, refine the codebook and reach consensus through deliberation. The final codes generated captured essential elements that occurred repeatedly throughout the dataset.
3. Generating Themes. Codes with thematic similarity were grouped together to form overarching themes, representing broader patterns and concepts that were significant in the context of the narratives provided and in relation to the study objectives.
4. Reviewing Themes. Themes generated in steps 1–3 above were presented to the Advisory Committee, along with other details of the coding process. Committee members reviewed the themes by checking them against the data to ensure they accurately reflect the material present in the data set.

### Findings

Seven themes were generated, listed in Table 2.

**Table 2:** Thematic findings from interviews and focus groups with patients and providers.

| Theme |
| --- |
| 1.Lack of voluntary, community-based mental healthcare within the context of the social determinants of health leads to involuntary psychiatric admissions and complicates discharges |
| 2.Involuntary psychiatric admission and treatment is a stressful experience for both people receiving and providing treatment and can lead to harm, trauma, and distrust in the healthcare system |
| 3.Peer support improves experience of care |
| 4.An “all or nothing” consent approach reduces patient autonomy and restricts rights |
| 5.Available treatments during involuntary admission are limited to a narrow range of options |
| 6.Stigma and discrimination occur |
| 7.The lack of conclusive evidence for involuntary treatment creates distress for care providers |

Theme 1: Lack of voluntary, community-based mental healthcare within the context of the social determinants of health leads to involuntary psychiatric admissions and complicates discharges Among the most salient themes found in both the interview and focus group data was the view that voluntary treatment, both in community to prevent people from reaching a state of mental health crisis and in hospital for those requiring acute care, is extremely limited and inaccessible to many. PWLLE and clinicians emphasized that community-based mental health supports are insufficient to meet demand, with programs often unavailable and unaffordable, and long wait lists and strict entry requirements making it hard to qualify for services.

> We shouldn’t have to prove, actively prove in front of them, that the situation is bad enough to actually get treatment. – PWLLE
>
> We don’t have enough acute psychiatric resources in the system, and we also don’t have enough at the other end. We don’t have enough places for people that have serious mental health dual diagnosis issues who don’t need to be in hospital but don’t have a place to go. – Clinician

The perspective of PWLLE and clinicians was that available mental health supports provided in community are insufficient to meet needs of people in this geographic region. This contributes to circumstances in which people reach a state of mental health crisis and perpetuates reliance on acute care. Both groups of participants assessed that, from a pragmatic perspective, the only meaningful way for many people to receive timely access to mental healthcare is to be admitted involuntarily to hospital. Interview participants reported that most people who receive psychiatric treatment in hospital are admitted involuntarily (with the exception of people receiving treatment for eating disorders, who participants observed tend to be predominantly treated voluntarily).

> People are uncomfortable with voluntary patients in an acute care setting … it almost feels like a luxury to be admitted voluntarily in an acute care setting. – Clinician

Even where preventative services are available, the stigma and ableism associated with “mental illness,” and the consequences of seeking help for such readily stigmatized conditions, were mentioned as barriers to accessing services. This in turn was reported to have negative implications for other determinants of health, such as employment.

> My employer has no idea that I have a mental illness. I cannot divulge. I would get degraded in my work, if not even let go – kind of pushed under the carpet – and one day I’m gone… So there needs a lot more done to also support people that are like me, living and hiding and have nowhere to run. – PWLLE

The role of social determinants of health — including housing, transportation, income, social support, food, spirituality, and culture — on mental wellness was also emphasized by PWLLE, clinicians, and other service providers. PWLLE reported their conditions are exacerbated by unmet basic needs, leading to mental health crises and repeat involuntary admissions.

Additionally, for those on extended leave, not having access to key social determinants of health can contribute to not being able to meet the conditions of extended leave (e.g., attending appointments), ultimately leading to recall to hospital.

> People go back to their lives that were the same as when they came in. – Clinician
>
> Somebody’s required to make an appointment and to show up on a day, or they’re getting recalled and going to the hospital. If that person is sleeping on the side of [the road], doesn’t have a clock, doesn’t know what day it is, doesn’t have the support to get there, and they missed their appointment with their mental health team. [Then] it’s negative documentation about their not following through, they’re not doing ABC and D. They’re getting recalled back to the hospital and they get put in same cycle again. – Clinician

Some clinicians noted that, in some cases, involuntary psychiatric admission and treatment was the only avenue for patients to access (or continue to access) resources to meet their basic needs. Clinicians frequently remarked that they felt “backed into a corner” to certify patients under the MH Act due to a lack of system-level support for voluntary care and community-based service coordination to address social determinants of health.

> I feel like that we get a little bit more backed into a corner to do involuntary treatment. Because I think we know that without it, sometimes this person might lose all these resources that they have, and it will be very hard to get them back. If our system was different, and people could get housing, and they could try again somewhere else, I think we would maybe feel less, or at least I would feel less, kind of stuck in the need for involuntary treatment in this context. – Clinician

Furthermore, clinicians described the challenges of working within an acute care setting focused on getting patients out of hospital quickly, with limited capacity to promote care continuity. The acute care system was seen by these participants as not being conducive to delivering quality care and supporting people post-discharge with access to supports promoting mental well-being. They also described the distress it caused them when they must discharge patients knowing there are few to no low-barrier, accessible resources to which to refer them.

> Usually, they give you a little slip of paper that says (sometimes not always), sometimes a slip of paper that says you can do this, that and the other thing. But you’re kind of left [on your own]. Sometimes it will say that you have a follow-up appointment but that only happened to me once. Otherwise, it’s just goodbye, good luck. And it’s terrifying going from the secure environment of a hospital back into the real world… especially if you live alone. It’s very scary. – PWLLE
>
> They just get discharged when they are good enough, you know, but they’re not. They’re just good enough to go home and not be taking up a $1,600 a day hospital bed. – Clinician

Clinicians described how a lack of resources in community even for those on extended leave can be a barrier to discharging a patient from hospital and can lead to patients having repeated episodes of involuntary psychiatric admission and treatment.

> Some of the more intensive mental health resources require that people have tried and failed different things. So, sometimes we know we’re setting someone out on extended leave, probably to a mental health service that’s not gonna meet their needs, and that it’s probably not gonna be successful and it’s going to lead to them being recalled, brought in by the police. – Clinician

Theme 2: Involuntary psychiatric admission and treatment is a stressful experience for both people receiving and providing treatment and can lead to harm, trauma, and distrust in the healthcare system

PWLLE described a spectrum of experiences with involuntary psychiatric treatment. Some indicated that, due to their mental state while in crisis, they would not have sought or accepted care voluntarily, and, looking back, were grateful to have received treatment.

> For me, at the onset of my illness, only outside intervention and getting me into hospital seems to work. I don’t think I would have actually survived, if I would have continued roaming on the street and not eating. – PWLLE

However, it was unclear whether the benefit was ascribed to the involuntary circumstances under which treatment was delivered, or if a similar outcome would have been achieved if some or all aspects of their care were voluntary.

> I am thankful that I did have involuntary treatment… I just wish there was a different process for going through it all like we’re sharing now, with options and stuff. – PWLLE

Some described a range of negative experiences during involuntary psychiatric admission. Participants repeatedly stated the use of coercive measures, including the understanding they were being “stripped of their rights” or being treated “like a criminal” when in hospital under the MH Act, led to fear and overwhelm, feeling stigmatized and traumatized. Some participants described feeling disempowered and being made to feel “like a child”.

> But if somebody like me gets stripped of all their rights and put into seclusion rooms — and close the door and injected and have no idea what’s going on — that’s a pretty brutal assault to one’s human dignity, to one’s human rights, to one’s perception of who we are in the community, how we are appreciated and included in a greater community. – PWLLE
>
> And I feel we are treated and under the law, we are [treated like a] criminal because our rights are taken away. We’re not criminals. We’re in a medical facility but we’re treated as a criminal that has done something wrong. – PWLLE
>
> You begin to feel like you’re being treated like a child when you’re actually an adult with a mental illness, which is very different than being a child. – PWLLE

PWLLE discussed how these negative experiences of involuntary psychiatric admission and treatment impacted them, including creating lasting trauma and lowered self-esteem. Some felt fearful of reliving the traumatic experience, which, in turn, discouraged them from seeking healthcare or fully disclosing their concerns to clinicians in the future. Health professionals corroborated this, noting they have witnessed ways that trauma associated with involuntary psychiatric admission and treatment can deter people from seeking care, both for mental and physical health concerns.

> Now for future, if I need to go back, I know I can’t be honest. I have to filter what I say or I’m going to be in a very scary situation that is going to, that is traumatic. And yeah, it’s taught me not to be honest when I seek mental healthcare. – PWLLE
>
> It also impacts access to healthcare because a lot of my clients will not bring themselves to the hospital or to a clinic if they’re having a health issue, because they’re afraid of being re-certified again. – Service Provider

Another consequence of involuntary psychiatric admission and treatment is that it can become a system barrier for patients to receive other forms of care and support, including accessing onsite programming at hospitals while admitted. For example, if a patient is in seclusion, they are unable to access other programs or services during that period.

The involvement of police in apprehending people under the MH Act was highlighted by PWLLE as a significant source of trauma, and by clinicians as compromising the therapeutic relationship. Experiencing, witnessing, or being aware of circumstances in which people experiencing mental health crises were “tasered”, “attacked”, handcuffed, or “dragged” by police while being detained and transported for involuntary treatment complicated and constrained participants’ willingness to engage with health services. For those participants, this created additional barriers to access. For some clinicians, police involvement created unnecessary complexities for healthcare delivery, creating additional strain on their ability to assess and care for patients in emergency settings.

> Since I was having an episode, so I decided to run instead of being safe and being like, to stand here. So, [the officer] attacked me, put me in cuffs. – PWLLE
>
> Separation between healthcare and police is super important… I don’t think police are trained or suited to make those [mental health] assessments. And in my experience, they’re the ones escalating the situation. – Clinician
>
> A lot of the times, the police will bring somebody, and they’ll say “look, we’re just told to bring them in. I have no idea why, like literally none.” I have to deal with it right? I’m responsible so those stress me out quite a bit. There’s no information whatsoever. The patient looks perfectly normal. There’s no supporting documentation why certain decisions got made. – Clinician

The majority of clinicians expressed a range of negative feelings related to involuntarily admitting and treating patients, including feeling burned out, traumatized, and inadequate due to working in a system that limits their ability to meet their patients’ needs. For instance, emergency department clinicians described the environment as unconducive to providing care to people experiencing a mental health crisis. The chaotic environment, lack of privacy, and lack of time to interact with patients limited their ability to deliver quality care.

> I think it is environment too. When you think about patients in Emergency on a stretcher being assessed and then involuntarily certified, you would just never want that for someone you care about. – Clinician
>
> I have to interview people in the parking lot, because every single care space is full, like literally. Everything is full. So, we have to go on the freezing parking lot. – Clinician

Clinicians felt hindered by not knowing a patient’s history and having limited time to observe a patient or establish a relationship. This was stated to increase the likelihood of activating “triggers” for PWLLE that may exacerbate mental health crises and determining a course of action that might not be of maximum benefit to the person they were treating.

> One of the things for example is the observation period. If the patient [within] two hours is completely rational, that’s an alteration period as well, where I’m going to be a lot less likely to do something. But if given only the five-second observation period to make decision, I’ll probably err on the side of certifying somebody. – Clinician

Some participants described adversarial feelings between clinicians and patients, between different clinicians, and between lawyers representing patients and clinicians, which can introduce additional risks and harms to all involved. For example, one participant commented:

> There are some [clinicians] that are great and have no problem with their patients, clients, whatever, applying for Review Panels. Others take that quite personally – that their judgment is being questioned – that makes it difficult. – Service Provider

Risks and fears discussed by clinicians included the conflicting fears that if they do not involuntarily detain someone, they will be responsible if “something bad happens”, and fears that if they do involuntarily detain someone, they could be held liable or sued. Clinicians also mentioned the risk of harm to staff by patients when attempting to deliver treatment against their patient’s will. Many participants talked about involuntary psychiatric admission and treatment being traumatic for patients, introducing additional emotional harms.

### Theme 3: Peer support improves experience of care

Many PWLLE felt having a peer to help them navigate the system while being admitted and treated would have been of tremendous benefit in allaying their fears and improving their experience.

> Another thing that would really, really help is peer support. Having [someone] there that could reassure me that, “OK this is what’s going to happen, you’re going to meet with the psychiatrist” or whatever the process is. And then if I’m admitted into the ward, or in this case certified, to explain what that means. – PWLLE

Clinicians agreed that peer support roles were valuable; however, they noted peer support workers are often treated as though they are not part of the care team with negative consequences for people in these roles.

> [Peers] had a really significant role in building relationships and helping people to access care. But the peers had a hell of a hard time, and we went through so many of them because they were stepping into an environment where there was fierce hierarchy. They didn’t even have letters after their names; they weren’t going to be taken seriously, and your self-esteem can only handle so much. – Clinician

Moreover, both PWLLE and interview participants felt including patient advisors with lived and living experience in planning services would provide valuable expertise for developing more holistic programs and services.

Another source of support identified by PWLLE was engagement and inclusion of family and friends. However, PWLLE had varied views regarding if and to what extent they would have liked their families to be informed about or involved in their involuntary psychiatric admission and treatment. Some participants felt family members could use more education on how to support their loved ones who are experiencing mental health crises. Others identified that family members, including chosen family and others providing support from within their communities, could serve as a valuable source of information to clinicians as they deliver care to people in crisis.

### Theme 4: An “all or nothing” consent approach reduces patient autonomy and restricts rights

PWLLE described a lack of control over all aspects of their treatment, and some reported they were not informed of what was happening — including not being informed of their rights under the MH Act. An “all or nothing” phenomenon seems to occur with the BC MH Act deemed consent approach, where once a person is involuntarily admitted under the MH Act, there is a default that all treatment and related interventions are delivered involuntarily. This approach leaves no room for patients to state their preferences or have those preferences considered in treatment planning.

> I think the hardest part about it was I felt like a persona non grata. I felt like nobody discussed anything with me. – PWLLE
>
> Nobody told me what was happening. Nobody told me that I had any rights, they just gave me two shots of Loxapine and I went to sleep. – PWLLE
>
> [I was told] “you’re here for shock treatment” and then they explained what that all meant. They did very good in that respect in explaining what shock treatment was, and even I got to watch a video about it. So that part was good. But no, there was no “Oh! by the way, do you want to get this?” – PWLLE

PWLLE described the importance of connection and compassion. Whenever they experienced empathy and compassion from clinicians or felt listened to and involved as much as possible in their care, this had a positive impact on their well-being and their engagement in care.

> Any time I’ve had a doctor or care provider that has included me in the decision-making or at least made it feel that way, I’ve found that the outcomes have been better and I’m more willing to take the medication. – PWLLE

They also noted that clinicians are very busy and burnt out, and it is challenging for staff to have enough time to meaningfully engage with their patients.

Some PWLLE emphasized information should be shared with patients in an easy-to-understand way and when they are more receptive.

> But the rights [under the MH Act] are so much, there is a lot of jargon, so it doesn’t really make sense to you, especially if you’re not in the right mindset to listen. – PWLLE

Participants also noted that when a person treated under the MH Act is informed of their rights, there are many process barriers to acting on this knowledge. For example, participants noted a potential for conflict when clinicians providing care are the ones to inform patients of their rights under the MH Act, including the right to a second medical opinion and a review panel. Moreover, they were concerned about the potential for a conflict of interest to arise when seeking second medical opinions, as the second physician consulted often works on the same care team as the first physician. In addition, there was concern there is no requirement for the first physician to change the treatment based on the second opinion. Concern was also expressed that when a person treated under the MH Act challenges their treatment at a review panel hearing, it is often a unit clerk who selects which health record documentation to provide to the lawyer representing the patient. In these circumstances, participants expressed concern that unit clerks may not have sufficient medical and legal knowledge to determine which parts of a patient chart are relevant. Another concern was that clinicians sometimes had to play roles during review panel hearings that impair their relationship with their patients.

> [It] creates a very weird dynamic, because they are acting kind of as counsel, they are also the expert witness; and, of course, they’re the client’s treating doctor. So, it can really impair the treatment relationship for it to proceed in that way. – Service Provider

### Theme 5: Available treatments are limited to a narrow range of options

When reflecting on options available for “treatment” to those being treated under the MH Act, medication, physical restraints, and seclusion were often mentioned. References to non-pharmaceutical treatment options, such as psychotherapy, tended to focus on how it is lacking. Some clinicians raised concerns about over-dependence on the use of physical and chemical restraints, as well as seclusion. A pertinent recommendation from one interview participant was to institute frequent re-assessments of patients in seclusion and hold debriefing sessions after every seclusion event to identify ways things could have been done differently. Clinicians and PWLLE alike expressed desire for meaningful and timely access to a much broader array of options, including counsellors and psychologists; recreation, music, and art therapy; and access to the outdoors.

> In emergency, people get thrown in seclusion because the nurses don’t have time, or they feel they don’t have time, or they’re worried that they’re going to abscond, or whatever, and they kind of lock them in there in an unethical way. But I don’t think that the nurses see it that way. They see it as they’re preserving their safety, whereas the patient probably wouldn’t feel that way. – Clinician
>
> There are so many things to do in response to someone yelling and only one of them is putting them in seclusion. – Service Provider

### Theme 6: Stigma and discrimination occur

Those who provide care and support to people who experience involuntary psychiatric admission and treatment noted some instances of discrimination, such as clinicians’ perceptions of risk and behaviour being influenced by racism (e.g., men of colour viewed as more dangerous) and sexism (e.g., women being judged on their appearance and sexual behaviour).

> I can tell you that I have unquestionably seen anti-Indigenous racism, sexism, ableism, operating in the attitudes of the people enforcing detention involuntary treatment which has changed the way the experience has played out for my clients. Yes, that’s what I think I could sort of confidently say. – Service Provider

There were also mentions of people being certified during pregnancy over concern for the fetus and then decertified once the baby was delivered. In addition, there were discussions of challenges for birthing parents who are involuntarily treated, as there is currently no option for them to have their newborns with them during an involuntary admission in these organizations’ facilities.

> There’re people who have been using [substances] and don’t necessarily have mental health challenges that I would assume would warrant certification in the hospital, but because they’re pregnant, that becomes the designated reason as to why they’re certified. – Service Provider

Examples of anti-Indigenous racism and discrimination against transgender people were also provided. For example, an interview participant shared that they had seen a patient chart that described an Indigenous patient who was speaking their language as speaking “gibberish.” As another example, interviewees described situations where a patient’s identification with a gender different than the one assigned to them at birth was viewed as a symptom of mental illness. In these examples, the lack of culturally safe or gender-affirming care is not only discriminatory, but clinicians were using patients’ behaviour in response to this discrimination as “evidence” of a mental health issue.

> The person recently is identifying as male and it says right in the report, the doctor’s handwritten notes, “This person likes to go by – the name that this person has chosen is Dave and the pronouns are he/him.” And it’s mentioned in handwritten notes a couple times that, “The client – patient – was upset and didn’t feel heard or understood.” And the physician referred to [the persons as] “her” consistently in handwritten notes, in the case note, and in the entire presentation. That’s not uncommon, that’s just one example I think of medicalizing and, “This person is delusional,” that kind of stuff. – Service Provider

Indigenous people serving in the role of liaisons for Indigenous patients in the health organizations were noted as providing culturally safe care and interview participants expressed a desire for people serving in this type of role. Also mentioned was the added layer of complexity in which Indigenous patients are held without their consent in a place where they have no access to cultural healing and wellness practices, language-speakers, or other tangible aspects of culturally-safe care.

There was concern that equity-oriented care and related principles — such as cultural safety, patient-centred care, harm reduction, gender-affirming, and trauma-informed practice — are buzzwords that are not translated into practice. Conscious, conspicuous, and transparent efforts are needed to ensure these principles are applied to practice within the context of involuntary psychiatric admission and treatment. This includes capacity for ongoing monitoring and evaluation focused on understanding how these efforts are experienced by care recipients and care providers, and what the tangible results are over time.

> I don’t think you can actually be trauma-informed when the policies and the system itself is just perpetuating that trauma. So, yeah, it feels a bit like an oxymoron. – Service Provider

### Theme 7: The lack of conclusive evidence for involuntary treatment creates distress for care providers

When considering the above issues together, and in light of their lived experience providing care, participants who have dedicated their careers to supporting people during mental health crises raised many questions and concerns about the lack of evidence for involuntary psychiatric admission and treatment. In so doing, they reflected openly on whether the presumed “benefits” of using the MH Act to intervene with someone experiencing a mental health crisis are substantiated by available evidence, and whether such approaches justify the harms they repeatedly observed occurring for their patients and clients when they are subjected to involuntary and coercive “care”. These participants expressed concern that little is known about the long-term impacts of removing a patient’s rights on their health and well-being, and about the financial impacts of continuing with “a carceral approach” to mental health service delivery at times when health systems are increasingly under pressure economically and related to adequate staffing levels.

For some, the experience of being asked to mandate care of unknown quality with unknown results was inconsistent with their goals to ensure all patients receive high-quality, evidence-based care. These participants advocated strongly for a transformation in how mental health crisis response is delivered, centring preventive, community-based, and voluntary supports that acknowledge and respond to social determinants of health and inequities in service access and treatment outcomes.

> Some of my concerns about this field, which seems unique to this field, is that the intention to help shuts down any evaluation about what the impact is… I’m not aware of any good evidence on evaluating what the long-term benefits or harms etc. are for involuntary intervention, so that’s something that feels problematic and unique to this area. – Service Provider
>
> When you mandate care that you know is good quality, then it’s one thing. If you mandate care that it’s unclear what the positive effect will be other than in the acute period, that’s very problematic or that brings out the more problematic aspect of mandated care which is terminating of freedom that a person has. – Clinician
>
> I know how expensive a carceral approach is, I know how much the police costs and I know how much having someone locked up in a psych ward cost. So yeah, I’d like to see a world where voluntary services are not only available (which doesn’t exist, so that’s a huge step), and then where the voluntary services are something that’s incentivized and that people feel like they want to go. – Clinician

In summary, our findings demonstrate that, overall, involuntary psychiatric admission and treatment under the MH Act is not experienced as consistent with the principles of equity-oriented care. This includes principles that care is experienced as safe, destigmatizing, trauma– and violence-informed, culturally-safe, harm reduction-oriented, anti-racist, and gender-affirming (25,26). PWLLE, clinicians and other professionals who support them, highlighted the stressful and often traumatic nature of involuntary psychiatric admissions and treatment. PWLLE discussed feeling afraid, overwhelmed, stigmatized, and disempowered. They described experiences of discrimination and environments not being conducive to providing care to people experiencing a mental health crisis. Concerns were also raised about treatment received being limited to primarily medication, restraints, and seclusion, and the insufficiency of being able to access the right to a second medical opinion and review panels. Moreover, the lack of voluntary, prevention-oriented, community-based mental health services and unmet basic needs exacerbate mental health issues, leading to a reliance on involuntary psychiatric admission and treatment. This lack of community-based services also made it challenging to provide adequate discharge planning. Practices that were experienced as helpful included peer support and instances where PWLLE felt listened to and involved as much as possible in their own care.

## Discussion

This work aimed to understand the extent to which two health organizations were or were not living up to their aspirations to deliver equity-oriented care that is experienced as safe, trauma– and violence-informed, culturally safe, harm-reduction oriented, anti-racist, and gender-affirming in the context of involuntary admission and treatment under the MH Act. Our findings demonstrate that, overall, although socialized and indicated in the language of some policies, these aspirations are not being achieved or fully implemented in practice. We identified many systemic factors mediating this outcome, as well as opportunities to move towards a more equity-focused approach in alignment with commitments to safeguarding human rights. We found that a lack of accessible voluntary, community-based mental healthcare, combined with unmet fundamental needs (such as housing, income, social support, food, spirituality, and culture), meant that mental health concerns are often not addressed until the point at which mental health crisis occurs. Involuntary psychiatric admission and treatment are often seen as the “only way” to access any mental healthcare.

Once an individual is admitted involuntarily, treatment options are limited and, by default, all psychiatric treatment and related interventions are delivered involuntarily. Involuntary psychiatric admission and treatment are stressful for both people receiving and providing treatment. Some participants experienced racism and transphobia during their involuntary psychiatric admission and treatment, and many reported experiencing stigma and discrimination.

Discharge planning from hospital is not supported or enabled by adequate resources in the community which contributes to the phenomenon of a “revolving door” where people are recertified numerous times in order to receive care or be connected to appropriate community resources. The negative experiences of involuntary psychiatric admission and treatment create lasting harm and discourage people from seeking healthcare in the future. There was also concern that human rights were being infringed upon, with little evidence of long-term effectiveness of involuntary psychiatric admission and treatment.

Our findings confirm what has been reported by other organizations and within the research literature. As in this project, research from other jurisdictions has found that in the context of involuntary psychiatric admission and treatment, people are subjected to chaotic and frightening care environments (44,45), coerced medication as a “taken-for-granted” practice (45), and being treated like children (15,45,46) or prisoners (44). Qualitative accounts elsewhere further affirm the perspectives of participants in this study: if they had been listened to and had their needs respected during the time of crisis, treatment could have occurred without coercive measures (45). In many cases, patients are not provided information about the reasons for their detention (15,46) or their rights (44,47) despite wanting this information (44). Authors elsewhere have also described the urgent need for advocates (15,44) and peer supports (44) to be available in hospital and community settings.

As in this study, previous research describes medication (15,44) and restraints (15) as being the main interventions used during involuntary psychiatric admission and treatment, with little use of other interventions that could be beneficial, such as psychotherapy and meaningful recreational, educational, or occupational activities (44,46). This literature contains a similar array of perceptions from people who have experienced involuntary psychiatric admission and treatment as were reported by the participants in our study.

While some studies have shown that people who have experienced involuntary psychiatric admission and treatment feel they have benefited from their treatment, and others documented perspectives that participants felt they would not have accepted treatment otherwise (15,46,48), others have expressed their treatment could have been provided through voluntary means if they had been given the opportunity for fully informed decision-making (15,44,48). Involuntary psychiatric admission and treatment has been described elsewhere in the literature as traumatizing (45,47), frightening (44), confusing (47), and stigmatizing (44,46), negatively impacting trust of healthcare providers (49), and leading people to avoid care in the future (45,50). Similarly, studies have found that the experience of providing involuntary psychiatric admission and treatment has a negative emotional and psychological impact on healthcare professionals (49).

Members of groups experiencing oppression and discrimination, such as Indigenous and racialized groups and young people, are impacted in numerous ways by coercive practices in mental healthcare (51). As highlighted by Morrow and Weisser (2012) “although involuntary detention of First Nations, Métis, Inuit and urban Indigenous children and youth under the MH Act may be intended for their safety and protection, it can be seen and experienced as another link in a long chain of oppression imposed by the state on Indigenous peoples” (p. 4). The “In Plain Sight” report similarly identified emergency departments and psychiatric services as settings in which Indigenous peoples in BC are subject to racist, discriminatory, and culturally-unsafe practices, including being detained involuntarily without clear rationale or for reasons informed by anti-Indigenous racism and stereotyping (53).

There is also ample literature attesting to the ways in which structural violence in the form of patriarchal, heteronormative, and transphobic practices have shaped, constrained, and undermined access to timely, appropriate, and supportive mental healthcare for women, gender-diverse people, and 2SLGBTQ+ communities, while normalizing violence and coercion in psychiatric treatment settings (54–57). Recent research has confirmed that gender-diverse and transgender people experience greater inequities in many dimensions of mental wellbeing, which are in turn often compounded by anti-transgender bias, non-affirmation, and further trauma when seeking mental healthcare (58,59). An Ontario study reported that expectations of negative or demeaning treatment in healthcare cause transgender people to avoid care to the extent that only 29 percent of transgender respondents seek healthcare for any reason (60). Of those who accessed emergency care, 52 percent reported experiencing “negative” (transphobic) treatment ranging from insulting or demeaning language to outright refusal of care (60).

Some interview participants in this study expressed concern there is limited evidence about whether involuntary treatment is effective, and this is also echoed in the literature. In a scoping review on ethical issues in clinical decision-making about involuntary psychiatric treatment, Laureano et al. (50) note that “the lack of data on the clinical benefit of involuntary hospitalization has been identified as a serious limitation” and from a systematic review on involuntary psychiatric hospitalization and long-term compliance, Cossu et al. (61) concluded that “although evidences carried out so far are weak, the data do not show a trend of improvements and do not seem to exclude the possibility of worse compliance after compulsory hospitalisation” (50). Similarly, prior systematic reviews found no evidence of patient benefits from community treatment orders (11).

Factors that contribute to involuntary psychiatric admission and treatment include a lack of preventative mental healthcare and wider social problems such as inadequate affordable housing, inequity, and discrimination (47). A lack of community-based resources to which to refer people after hospitalization was reported as a concern. [76] Things that contributed to the negative impacts of involuntary psychiatric admission and treatment included the dominance of “risk management” in healthcare (49), and insufficient time for clinicians to build relationships with people receiving psychiatric treatment. Experiences of racism and discrimination against those with low socio-education levels leading to of them being subjected to involuntary psychiatric admission and treatment at higher rates (50) were described in the literature. Participants in our study spoke of witnessing treatment of Indigenous patients that were aligned with the findings of the In Plain Sight report (62) which detailed that Indigenous people are subject to racist, discriminatory, and culturally unsafe practices. Similarly, clinician participants in our study spoke of treatment of transgender patients that aligned with previous studies showing that gender diverse and transgender people experience anti-transgender bias, non-affirmation, and further trauma when seeking mental healthcare (58,59).

The importance of clinicians listening to, understanding, and respecting patients (15,46) to build trust to improve the quality of care during psychiatric treatment (45) is a common theme in the literature on involuntary psychiatric treatment. Conversely, when clinicians do not talk to patients and allow them to explore their feelings, it can increase patient’s agitation and result in the use of medications and restraints (15). Patients suggested that making choices is important to their recovery and being involved in decision-making about their care could increase as their recovery progresses (44). Others have also documented similar recommendations to the ones arising from this project, including the critical importance of listening to patients, building trusting relationships, developing individualized and flexible care plans and discharge plans in cooperation with patients (45), eliminating anti-Indigenous racism and providing culturally safe care (63), and avoiding coercive measures (45,63). Moreover, there were calls for more early intervention to provide voluntary interventions before a crisis begins (50,63) and government funding to address underlying issues leading to mental health crisis, such as inadequate affordable housing, discrimination, and other social determinants of health (47,63).

Globally, movements for health justice and health equity are emphasizing the critical importance of aligning mental health service provision to international commitments to promote well-being and safeguard human rights (64,65). Canada is a signatory to many international agreements, including United Nations Conventions, which oblige Canadian provinces to align domestic legislation to remain in compliance. For instance, the 2006 United Nations Convention on the Rights of Persons with Disabilities (UN CRPD) requires signatory countries to undertake substantive reforms to protect and promote human rights in mental health services, and to ensure a continuum of community-based supports are available to prevent conditions which result in institutionalization of all kinds. Article 19 of the UN CRPD requires signatory States to recognize “the equal right of all persons with disabilities to live in the community, with choices equal to others” and to “take effective and appropriate measures to facilitate full enjoyment by persons with disabilities of this right” (66). This includes the right to “have access to a range of in-home, residential and other community support services, including personal assistance necessary to support living and inclusion in the community, and to prevent isolation or segregation from the community” (66). As noted by the UN Special Rapporteur on the Rights of Persons with Disabilities in her End of Mission Statement following her 2019 visit to Canada, involuntary admission and treatment in BC are in contradiction to articles 14 and 25 of the CRPD, including “absence of procedural guarantees, the lack of alternative treatment options” and insufficient monitoring of designated facilities (67). She further noted that there is a “lack of comprehensive responses to guarantee the access of persons with disabilities to the support they need to live independently in their communities” (67). Our findings confirm this. The UN Special Rapporteur also called on governments to provide support, including adequate housing, and for the meaningful participation of people with disabilities in decision-making, which aligns with our recommendations.

The guiding principles for BC’s mental health law and services developed by Health Justice (25), advocate for efforts to bring the BC Mental Health Act “up to date with evidence-based best practices and human rights principles” (68). These principles note the importance of recognizing human rights, using a holistic approach to mental wellness, providing high-quality health services, complying with the UN Declaration on the Rights of Indigenous Peoples, prioritizing intersectional equity, supporting self-determination, including the meaningful involvement of people with lived and living experience of involuntary psychiatric admission and treatment in all aspects of decision-making that impacts them. They also call for more accountability and oversight considering the extraordinary power of the MH Act over people’s human rights. The findings of this study emphasize the urgent need to align mental healthcare delivery with these guiding principles.

## Strengths and Limitations

This project was guided by an Advisory Committee comprised of individuals with lived and living experience of receiving or providing involuntary psychiatric admission and treatment, service providers in community agencies supporting those receiving care under the MH Act, clinicians and leaders working with the health organizations’ mental health and substance use services, and researchers with expertise in equity-oriented mental health policy and practice. We used a trauma-informed approach to the data collection and focus groups for PWLLE were led by PWLLE of involuntary psychiatric admission and treatment.

Following the Committee’s advice, we recruited participants for the focus groups for PWLLE through both social media and through community agencies that work with PWLLE. Interested participants were later screened to exclude those who did not receive involuntary treatment in the specific health organizations in which the study was situated. We did not recruit directly through the health organizations or their services directly. As part of our recruitment process, and to ensure transparency, we disclosed that this study was commissioned by the health organizations, and that results would be communicated directly to it. Consequently, we may have not reached those experiencing the greatest structural vulnerabilities when interacting with health systems, or those whose experiences of trauma during their interactions with health systems would have discouraged them from participating in a study involving those health organizations.

This is a study with a small sample size using qualitative methods, and, as is typical with qualitative research of this type, results may not be generalizable to other contexts or representative of all patient or provider experiences or perspectives. While the interview guide and recruitment materials were neutral with respect to the type of experiences sought, the overwhelming majority of participants in both the treatment recipient and the care provider groups (clinician and service providers) spoke of negative, stressful, or concerning experiences. No participants spoke of experiences that were primarily positive. As a result, this study is not able to comment on policies, practices, or contexts which participants associated with positive care experiences. There was a high rate (66%) of people who registered for the in-person focus groups who did not attend the focus group; however, this did not occur in virtual focus groups, in which all who registered attended. In addition, 17 PWLLE participants did not disclose their age, gender, or other demographic information, limiting opportunities for intersectional analyses of these experiences.

## Conclusions

As is demonstrated by the above findings and supported by an ample body of evidence in other settings and jurisdictions, compliance with the administrative requirements of the MH Act alone as encouraged by BC’s Ombudsperson’s report (8) does not guarantee quality of care, alignment with commitments to equity-oriented care, or the protection of human rights. The delivery of evidence-based mental healthcare in all forms requires health system and service planners to acknowledge mounting evidence within the international context that insists upon urgent movement away from coercive care practices. People with lived and living experience of receiving and providing involuntary admission and treatment in this BC health authority identified a serious and concerning lack of system-level supports and pathways for prioritizing and enabling low-barrier access to voluntary, community-based mental health supports and for addressing the social determinants of health that often precipitate mental health crises.

## Data Availability

The data supporting this study are not publicly available due to participant privacy concerns. Researchers wishing to access the full data set should contact to request access, outlining their intended use of the data.

## Acknowledgements

The authors would like to acknowledge all members of the advisory committee who provided valuable guidance on all aspects of the study.

